# Prevalence of bacterial pathogens and potential role in COVID-19 severity in patients admitted to intensive care units in Brazil

**DOI:** 10.1101/2020.12.22.20248501

**Authors:** Fabíola Marques de Carvalho, Leandro Nascimento Lemos, Luciane Prioli Ciapina, Rennan Garcias Moreira, Alexandra Gerber, Ana Paula C Guimarães, Tatiani Fereguetti, Virgínia Antunes de Andrade Zambelli, Renata Avila, Tailah Bernardo de Almeida, Jheimson da Silva Lima, Shana Priscila Coutinho Barroso, Mauro Martins Teixeira, Renan Pedra Souza, Cynthia Chester Cardoso, Renato Santana Aguiar, Ana Tereza Ribeiro de Vasconcelos

## Abstract

Secondary bacterial and fungal infections are associated with respiratory viral infections and invasive mechanical ventilation. In Coronavirus disease 2019 (COVID-19), lung injury by SARS-CoV-2 and impaired immune response can provide a favorable environment for microorganism growth and colonization in hospitalized individuals. Recent studies suggest that secondary bacterial pneumonia is a risk factor associated with COVID-19. In Brazil, knowledge about microbiota present in COVID-19 patients is incipient. This work describes the microbiota of 21 COVID-19 patients admitted to intensive care units from two Brazilian centers. We identified respiratory, nosocomial and bacterial pathogens as prevalent microorganisms. Other bacterial opportunistic and commensal species are also represented. Virulence factors of these pathogenic species, metabolic pathways used to evade and modulate immunological processes and the interconnection between bacterial presence and virulence in COVID-19 progression are discussed.

**Article Summary Line:** We identified respiratory, nosocomial and bacterial pathogens as prevalent microorganisms in 21 Brazilian COVID-19 patients admitted to Intensive Care Units. Pathogen virulence factors and immune response evasion metabolic pathways are correlated to COVID-19 severity.

## Introduction

Severe COVID-19 cases are characterized by systemic hyperinflammation and immune dysfunction, which may lead to lung damage. Infection begins with the internalization of the viral complex through the interaction between SARS-CoV-2 spike protein and the angiotensin-converting enzyme 2 (ACE2) in the host cell. ACE2 expression reduces after SARS-CoV-2-infection, resulting in an increase in angiotensin II which lead to pathophysiological effects that include vasoconstriction, increased inflammation and fibrosis (*1*). Lung edema, endothelial and epithelial injuries are accompanied by an influx of neutrophils into the interstitium and bronchoalveolar space, resulting in impairment of arterial oxygenation (*2*). Infection of alveolar macrophages by SARS-CoV and neutrophil infiltration are important triggers of the cytokine storm, which is extensively associated with COVID-19 severity and mortality (*3*).

In the critical stage of COVID-19, cytokine storm is continuous, contributing to vascular congestion, complement cascade activation and over disseminated intravascular coagulation (*4*). Hypoxia is aggravated and invasive mechanical ventilation may be required for life maintenance. Under these conditions, variation in oxygen tension, alveolar ventilation, deposition of inhaled particles, blood flow and concentration of inflammatory cells are factors that directly affect lung microbial growth conditions (*5, 6*). In addition, irregular innate immune response in acute lung injury, mainly during the process of neutrophil recruitment, may favor bacterial infections, which are also regulated by virulence factors (*7*).

In Influenza and Severe Acute Respiratory Syndrome (SARS), secondary bacterial infection has been reported during intensive care unit admission and during use of invasive mechanical ventilation. Data in literature pointed out that occurrence of co-infection or secondary bacterial pneumonia was observed in 11% to 35% of individuals with laboratory confirmed influenza (*8*), and that *Streptococcus pneumoniae* and *Staphylococcus aureus* were present in 35% and 28% of patients, respectively. In Canada, *Chlamydophila pneumoniae* or *Mycoplasma pneumoniae* were abundant in up to 30% of SARS patients (*9*). Bacterial infection was also identified in COVID-19 cases (*10 - 12*). Studies conducted in China, the USA, Spain and Thailand, showed that secondary bacterial infection was present in 14% of patients, with occurrence of *Streptococcus pneumoniae, Klebsiella pneumoniae* and *Haemophilus influenzae* within 1–4 days of COVID-19 disease onset (*13,14*). According to Rawson et al. (2020)(*15*), the extensive use of broad-spectrum antibiotics can predispose COVID-19 patients to acquisition of bacterial infections and increase multidrug-resistance.

Colonizing species in patients infected with SARS-CoV-2, prevalence of bacterial infection and mechanism of co-or secondary-infection remain poorly understood. In Brazil, an investigation of bacterial microbiota in SARS-CoV-2 was conducted in only one patient and concise results showed the predominance of *Lautropia, Prevotella* and *Haemophilus* genera (*16*). In-depth work was not performed.

This study aims to determine the microbiota of 21 COVID-19 patients admitted to intensive care units from two Brazilian centers, focusing on prevalence of bacterial pathogens, and to correlate the microbiota with the immunological disorder characteristic of Coronavirus disease 2019.

## Material and Methods

### Study cohort and data collection

Twenty-one COVID-19 patients admitted to Intensive Care Units (ICU) from Hospital Naval Marcílio Dias (Rio de Janeiro) and Hospital Eduardo de Menezes (Belo Horizonte) were included in this study. Both hospitals are located in cities in the Southeast region of Brazil. All patients had positive RT-PCR tests for SARS-CoV-2 (Table 1). Samples identification (IDs) were modified to preserve the patient’s integrity. The study cohort included 12 males and 9 females and no age restrictions were applied (ranging from 37 to 89 years of age).The present study was approved by the Ethics Commission from Hospital Naval Marcílio Dias and Hospital Eduardo de Menezes (protocol number 32382820.3.0000.5256 and 31462820.3.0000.5149). Clinical and demographic characteristics of the 21 patients were obtained from medical records and summarized in Table1.

**Table 1.** Summary of the clinical and demographic aspects of the 21 COVID-19 patients analyzed.

| Samples ID* | CT RT-PCR - N1 | CT RT-PCR - N2 | Intervals (first symptom sampling) (days) | Age range (years) | Sex | Risk factor | Ventilation use | X-ray | Chest computed tomography | Hospitalization (days) | Death |
| --- | --- | --- | --- | --- | --- | --- | --- | --- | --- | --- | --- |
| BH1 | 22.36 | 28.23 | 4 | 80 -90 | Male | Obesity; advanced age | Yes; invasive | Mixed | Not performed | 28 | Yes |
| BH2 | 34.75 | 26.81 | 5 | 80 -90 | Female | Advanced age | Yes; invasive | Interstitial infiltrate | Not performed | 6 | Yes |
| BH3 | 26.52 | 29.57 | 5 | 50-60 | Male | Immunocompromised | Yes; invasive | Interstitial infiltrate | Not performed | 55 | No |
| BH4 | 26.8 | 29.3 | 5 | 50-60 | Female | Diabetes; obesity | Yes; invasive | Interstitial infiltrate | Not performed | 23 | No |
| BH5 | 20.2 | 19.92 | 6 | 40-50 | Male | Diabetes; hypertension | Yes; invasive | Interstitial infiltrate | Not performed | 7 | Yes |
| BH6 | 33.59 | 37.28 | 6 | 80 -90 | Male | Advanced age | Yes; invasive | Interstitial infiltrate | Not performed | 11 | Yes |
| BH7 | 33.89 | 37.66 | 7 | 40-50 | Male | Diabetes; hypertension; ex-smoker | Yes; invasive | Interstitial infiltrate | Not performed | 60 | Yes |
| BH8 | 29.2 | 30.6 | 7 | 30-40 | Male | Diabetes | Yes; invasive | Normal | Not performed | 3 | Yes |
| BH9 | 19.7 | 25.3 | 7 | 60-70 | Male | Diabetes; obesity; hypertension; advanced age | Yes; invasive | Interstitial infiltrate | Not performed | 2 | Yes |
| BH10 | 17.98 | 18.97 | 7 | 40-50 | Female | Obesity | Yes; invasive | Consolidation | Not performed | 7 | Yes |
| BH11 | 18.63 | 20.73 | 8 | 40-50 | Female | Diabetes; chronic heart disease | Yes; invasive | Interstitial infiltrate | Not performed | 10 | Yes |
| BH12 | 25.46 | 26.56 | 8 | 50-60 | Male | Retocolitis | Yes; not invasive | Mixed | Not performed | 17 | No |
| BH13 | 27.87 | 29.36 | 9 | 60-70 | Female | COPD; diabetes; obesity; hypertension | Yes; invasive | Interstitial infiltrate | Not performed | 10 | Yes |
| BH14 | 16.94 | 18.15 | 10 | 50-60 | Female | Obesity; chronic heart disease | Yes; invasive | Interstitial infiltrate | Not performed | 13 | Yes |
| RJ1 | 24 | 26 | 11 | 50-60 | Male | Obesity; hypertension | Yes; invasive | Not performed | COVID suggestive with WITH 50-75% damage | 17 | No |
| BH15 | 26 | 29.5 | 12 | 60-70 | Male | Diabetes; chronic heart disease; advanced age | Yes; invasive | Not performed | Not performed | 18 | Yes |
| BH16 | 35.56 | 32.73 | 15 | 70-80 | Male | Diabetes; chronic heart disease; advanced age | Yes; invasive | Not performed | Not performed | 2 | Yes |
| BH17 | 15.9 | 16.3 | 16 | 60-70 | Female | Hypertension; advanced age | Yes; invasive | Mixed | Not performed | 6 | Yes |
| BH18 | 24 | 28 | 16 | 80 -90 | Female | Chronic heart disease; advanced age | Yes; invasive | Interstitial infiltrate | Not performed | 10 | No |
| RJ2 | 32 | 32 | 18 | 50-60 | Male | None | Yes; invasive | Not performed | Ground-glass images and consolidation areas with impairment of about 25-50% of the lung parenchyma | 20 | No |
| RJ3 | 30 | 32 | 34 | 70-80 | Female | Diabetes; hypertension; advanced age | Yes; invasive | Not performed | Pulmonary damage >50% | 37 | Yes |

\* Sample IDs were modified to preserve the patient's integrity.

### DNA extraction and sequencing

DNA was extracted using standard manufacturer’s protocols for the Qiagen QIAamp DNA Microbiome kit (Qiagen, Germany). Metagenomic libraries were constructed using the Nextera DNA Flex Library Preparation Kit (Illumina, USA). Sequencing was performed in a NextSeq 500 System with a NextSeq 500/550 High Output Kit v2.5 (300 Cycles) (Illumina, USA).

### Bioinformatics analysis

Sequencing files were submitted to MG-RAST (version 4.0.3) (*17)* for taxonomical and functional inferences. Sequences were trimmed using default parameters and human sequences removed by screening against *Homo sapiens*. Species were identified from MG-RAST using RefSeq and functional abundances were obtained using SEED subsystem database. For both analyses, the Best Hit Classification criteria and alignment cutoff applied included an e-value > 10^*-5*^, a minimum identity of 60%, and a minimum alignment of 15. Low abundance species (≤ 50 absolute counts of the sum of all samples) were removed to avoid false-positives and minimize the error of taxonomical predictions (*18*). The number of species was resampled so that all samples had the same library size. Prevalent species were detected using the microbiome R package (*19*), considering only those that occurred in 75% of all patients. Phyloseq R package was used to estimate the Shannon diversity index (*20*).

## Results and Discussion

The study included only adult patients, the majority of whom were above 65 years of age (42.8%). Males represent 57% of the patients sampled. The period between symptom onset and sample collection varied from 4 to 34 days. All individuals were admitted to intensive care units and invasive ventilatory support was necessary in 95.2% of the cases. No previous pulmonary impairment was reported, except for one individual. Most patients had at least one of the main comorbidities associated with disease severity. Diabetes represented 50% of the cases studied, followed by obesity and hypertension (33% in both) and cardiovascular disease (23.8%) (Table 1). Tracheal lavage samples with SARS-CoV-2 infectivity had higher abundances of bacterial respiratory and nosocomial species (Table 2). To check the relationship between microbial diversity and the proportion of microbial pathogens, we estimated diversity using the Shannon index and calculated the relative abundance of pathogens (Figure 1). The proportion of microbial pathogens was based on the relative abundance of individual pathogens divided by the total number of species in the microbiome. First, we observed a negative correlation (−0.9733005; p ≤ 0.05) between these two parameters, indicating that the prevalence of pathogens in all samples is inverse to diversity, causing a decrease in species richness. A high proportion of pathogens can be explained by the potential imbalance between microbial immigration and elimination, and regional growth conditions, as proposed by Dickson and collaborators (2015)(*21*).

**Table 2.** Abundance of the fifty most prevalent species obtained from COVID-19 patients using MG-RAST inference.

| Species | BH1 | BH2 | BH3 | BH4 | BH5 | BH6 | BH7 | BH8 | BH9 | BH10 | BH11 | BH12 | BH13 | BH14 | RJ1 | BH15 | BH16 | BH17 | BH18 | RJ2 | RJ3 |
| --- | --- | --- | --- | --- | --- | --- | --- | --- | --- | --- | --- | --- | --- | --- | --- | --- | --- | --- | --- | --- | --- |
| *Acinetobacter baumannii* | 150 | 78 | 460 | 5 | 31 | 91 | 462 | 100 | 161 | 6 | 65 | 9 | 78 | 1443 | 23845 | 10 | 97 | 482 | 19 | 23949 | 24061 |
| *Streptococcus pneumoniae* | 425 | 177 | 357 | 978 | 14121 | 43 | 577 | 11 | 82 | 4212 | 5965 | 2484 | 67 | 2563 | 0 | 1925 | 45 | 154 | 6364 | 0 | 0 |
| *Pseudomonas aeruginosa* | 93 | 524 | 225 | 8 | 8 | 58 | 237 | 13750 | 199 | 389 | 24 | 8 | 311 | 54 | 23 | 41 | 285 | 303 | 7 | 44 | 34 |
| *Staphylococcus aureus* | 498 | 5520 | 365 | 152 | 81 | 4643 | 263 | 29 | 281 | 352 | 210 | 278 | 138 | 720 | 0 | 1653 | 181 | 209 | 142 | 0 | 0 |
| *Enterococcus faecalis* | 197 | 401 | 233 | 535 | 620 | 174 | 343 | 16 | 137 | 460 | 497 | 1326 | 78 | 343 | 0 | 589 | 128 | 175 | 778 | 0 | 0 |
| *Escherichia coli* | 85 | 139 | 161 | 27 | 41 | 31 | 165 | 96 | 232 | 32 | 167 | 55 | 216 | 140 | 74 | 59 | 206 | 192 | 65 | 104 | 102 |
| *Listeria monocytogenes* | 220 | 79 | 275 | 66 | 72 | 141 | 197 | 6 | 376 | 51 | 124 | 135 | 110 | 124 | 0 | 94 | 246 | 232 | 98 | 0 | 0 |
| *Streptococcus agalactiae* | 35 | 17 | 42 | 117 | 140 | 4 | 61 | 3 | 11 | 307 | 120 | 117 | 16 | 63 | 0 | 57 | 10 | 29 | 327 | 0 | 0 |
| *Streptococcus pyogenes* | 44 | 26 | 24 | 99 | 137 | 9 | 50 | 5 | 11 | 285 | 116 | 107 | 26 | 63 | 1 | 59 | 4 | 12 | 364 | 0 | 0 |
| *Streptococcus spp.* | 6 | 2 | 12 | 15 | 515 | 1 | 12 | 0 | 1 | 126 | 246 | 41 | 1 | 47 | 0 | 68 | 0 | 5 | 149 | 0 | 0 |
| *Acinetobacter spp. ADP1* | 3 | 5 | 32 | 2 | 3 | 2 | 39 | 8 | 20 | 0 | 3 | 1 | 11 | 34 | 350 | 2 | 9 | 40 | 1 | 390 | 394 |
| *Bacillus cereus* | 555 | 152 | 890 | 38 | 86 | 349 | 806 | 28 | 1398 | 72 | 245 | 79 | 374 | 372 | 2 | 107 | 872 | 900 | 72 | 1 | 2 |
| *Bacillus anthracis* | 481 | 136 | 728 | 40 | 72 | 241 | 630 | 35 | 1228 | 53 | 175 | 66 | 329 | 286 | 0 | 61 | 691 | 673 | 72 | 0 | 0 |
| *Bacillus thuringiensis* | 197 | 46 | 304 | 13 | 27 | 130 | 326 | 9 | 549 | 16 | 92 | 32 | 145 | 137 | 0 | 34 | 348 | 333 | 38 | 0 | 0 |
| *Burkholderia pseudomallei* | 59 | 168 | 251 | 4 | 28 | 39 | 228 | 25 | 241 | 16 | 28 | 10 | 351 | 154 | 10 | 50 | 318 | 316 | 4 | 9 | 4 |
| *Streptococcus suis* | 56 | 35 | 50 | 121 | 155 | 7 | 69 | 42 | 31 | 370 | 167 | 159 | 32 | 134 | 0 | 96 | 18 | 39 | 368 | 0 | 0 |
| *Acidovorax spp.* | 144 | 877 | 787 | 2 | 9 | 50 | 965 | 18 | 254 | 28 | 25 | 3 | 1522 | 89 | 1 | 160 | 1073 | 1002 | 5 | 0 | 6 |
| *Comamonas testosteroni* | 8 | 231 | 221 | 0 | 2 | 7 | 292 | 7 | 65 | 11 | 6 | 1 | 493 | 18 | 1 | 46 | 348 | 334 | 2 | 0 | 0 |
| *Cupriavidus metallidurans* | 33 | 140 | 129 | 2 | 4 | 11 | 146 | 23 | 75 | 3 | 16 | 1 | 239 | 32 | 1 | 26 | 193 | 204 | 3 | 2 | 1 |
| *Streptococcus mitis* | 61 | 17 | 50 | 145 | 1571 | 6 | 56 | 0 | 12 | 358 | 646 | 236 | 4 | 263 | 0 | 194 | 4 | 18 | 717 | 0 | 0 |
| *Pseudomonas putida* | 16 | 2727 | 64 | 3 | 6 | 15 | 60 | 124 | 24 | 4 | 8 | 6 | 91 | 12 | 3 | 15 | 86 | 80 | 4 | 12 | 10 |
| *Stenotrophomonas maltophilia* | 380 | 81 | 78 | 0 | 8 | 84 | 61 | 9 | 630 | 15 | 43 | 17 | 122 | 328 | 4 | 37 | 381 | 77 | 17 | 24 | 1 |
| *Delftia acidovorans* | 35 | 3895 | 3129 | 0 | 2 | 13 | 3868 | 2 | 67 | 101 | 12 | 1 | 6808 | 23 | 0 | 693 | 4336 | 4359 | 2 | 3 | 0 |
| *Coprobacillus spp.* | 3061 | 322 | 414 | 113 | 206 | 232 | 158 | 198 | 640 | 3 | 1335 | 24 | 471 | 829 | 260 | 42 | 297 | 103 | 420 | 2 | 3 |
| *Enterococcus faecium* | 271 | 201 | 167 | 155 | 241 | 119 | 227 | 1 | 196 | 188 | 276 | 322 | 65 | 211 | 0 | 206 | 146 | 151 | 327 | 0 | 0 |
| *Staphylococcus epidermidis* | 129 | 54 | 126 | 42 | 12 | 12147 | 42 | 2 | 93 | 328 | 23 | 439 | 56 | 200 | 0 | 4022 | 40 | 50 | 26 | 0 | 0 |
| *Bifidobacterium dentium* | 20 | 12 | 456 | 11 | 3 | 1 | 10 | 0 | 0 | 4 | 9 | 457 | 17 | 584 | 0 | 8 | 2 | 26 | 7 | 0 | 0 |
| *Clostridium difficile* | 403 | 16 | 109 | 92 | 67 | 16 | 123 | 7 | 74 | 1802 | 642 | 141 | 77 | 133 | 0 | 151 | 44 | 39 | 126 | 1 | 0 |
| *Granulicatella adiacens* | 19 | 265 | 8 | 261 | 289 | 1 | 39 | 1 | 2 | 44 | 164 | 396 | 2 | 106 | 0 | 145 | 3 | 10 | 512 | 0 | 0 |
| *Streptococcus salivarius* | 8 | 1 | 10 | 486 | 111 | 8 | 46 | 0 | 1 | 57 | 137 | 126 | 22 | 40 | 0 | 46 | 0 | 2 | 1611 | 0 | 0 |
| *Streptococcus anginosus* | 37 | 1 | 36 | 12 | 12 | 2 | 80 | 0 | 1 | 2124 | 200 | 15 | 12 | 35 | 0 | 8 | 8 | 0 | 33 | 0 | 0 |
| *Rothia mucilaginosa* | 44 | 1 | 134 | 16 | 131 | 1 | 44 | 1 | 0 | 13 | 58 | 46 | 2 | 847 | 0 | 3963 | 0 | 6 | 662 | 0 | 0 |
| *Atopobium parvulum* | 156 | 30 | 58 | 1193 | 36 | 1 | 44 | 0 | 5 | 246 | 332 | 768 | 36 | 392 | 0 | 1009 | 1 | 271 | 45 | 0 | 0 |
| *Actinomyces odontolyticus* | 70 | 4 | 38 | 120 | 39 | 11 | 189 | 0 | 3 | 12 | 140 | 2744 | 14 | 226 | 0 | 684 | 0 | 58 | 43 | 0 | 0 |
| *Streptococcus parasanguinis* | 57 | 25 | 73 | 334 | 346 | 2 | 103 | 0 | 0 | 282 | 212 | 498 | 1 | 508 | 0 | 180 | 3 | 88 | 1629 | 0 | 0 |
| *Parvimonas micra* | 338 | 10 | 15 | 84 | 4 | 2 | 119 | 0 | 0 | 1309 | 747 | 36 | 284 | 196 | 0 | 13 | 6 | 1 | 21 | 0 | 0 |
| *Streptococcus infantis* | 16 | 6 | 2 | 14 | 1315 | 2 | 7 | 1 | 0 | 85 | 388 | 61 | 1 | 72 | 0 | 155 | 1 | 2 | 271 | 0 | 0 |
| *Streptococcus sanguinis* | 29 | 12 | 61 | 192 | 242 | 2 | 143 | 1 | 0 | 629 | 210 | 118 | 8 | 108 | 0 | 79 | 4 | 18 | 384 | 0 | 0 |
| *Streptococcus gordonii* | 47 | 17 | 72 | 94 | 241 | 3 | 103 | 0 | 2 | 684 | 219 | 124 | 10 | 159 | 0 | 66 | 3 | 15 | 359 | 0 | 0 |
| *Leptotrichia buccalis* | 36 | 2 | 19 | 9 | 8 | 8 | 6 | 4 | 0 | 15 | 7 | 1202 | 9 | 6 | 0 | 64 | 0 | 1 | 607 | 0 | 0 |
| *Veillonella parvula* | 39 | 1 | 10 | 222 | 36 | 22 | 39 | 2 | 4 | 18 | 62 | 321 | 64 | 63 | 0 | 338 | 1 | 12 | 69 | 0 | 0 |
| *Propionibacterium acnes* | 232 | 26 | 51 | 14 | 246 | 189 | 71 | 2 | 108 | 13 | 47 | 199 | 92 | 206 | 2 | 104 | 38 | 133 | 13 | 0 | 0 |
| *Bacillus subtilis* | 730 | 179 | 963 | 52 | 77 | 393 | 872 | 28 | 1361 | 53 | 221 | 61 | 378 | 480 | 5 | 77 | 995 | 1103 | 75 | 0 | 1 |
| *Streptococcus thermophilus* | 24 | 15 | 39 | 704 | 137 | 6 | 91 | 2 | 6 | 150 | 216 | 223 | 35 | 106 | 0 | 66 | 3 | 12 | 2129 | 0 | 0 |
| *Bifidobacterium longum* | 26 | 37 | 143 | 68 | 11 | 7 | 22 | 0 | 3 | 17 | 27 | 181 | 944 | 196 | 0 | 127 | 3 | 513 | 68 | 0 | 0 |
| *Lactobacillus gasseri* | 21 | 10 | 3 | 1615 | 3 | 5 | 5 | 0 | 11 | 107 | 6 | 32 | 4 | 2 | 0 | 7 | 6 | 2 | 14 | 0 | 0 |
| *Bacillus coagulans* | 179 | 42 | 181 | 1 | 15 | 86 | 152 | 0 | 329 | 2 | 42 | 4 | 71 | 115 | 1 | 14 | 246 | 185 | 5 | 0 | 0 |
| *Acidovorax ebreus* | 46 | 130 | 142 | 1 | 5 | 18 | 146 | 8 | 84 | 4 | 12 | 1 | 232 | 22 | 0 | 28 | 160 | 186 | 3 | 2 | 1 |
| *Candida albicans* | 29580 | 1104202 | 1136 | 21439 | 53 | 1407860 | 0 | 14 | 1 | 248985 | 8 | 43 | 18 | 72 | 30 | 66628 | 6 | 1176 | 3 | 203 | 179701 |
| *Candida tropicalis* | 307 | 803954 | 6950 | 189 | 3 | 12343 | 3 | 2 | 0 | 2194 | 1 | 19 | 3 | 0 | 1 | 556 | 4 | 2786 | 0 | 6 | 1526 |

**Figure 1.**
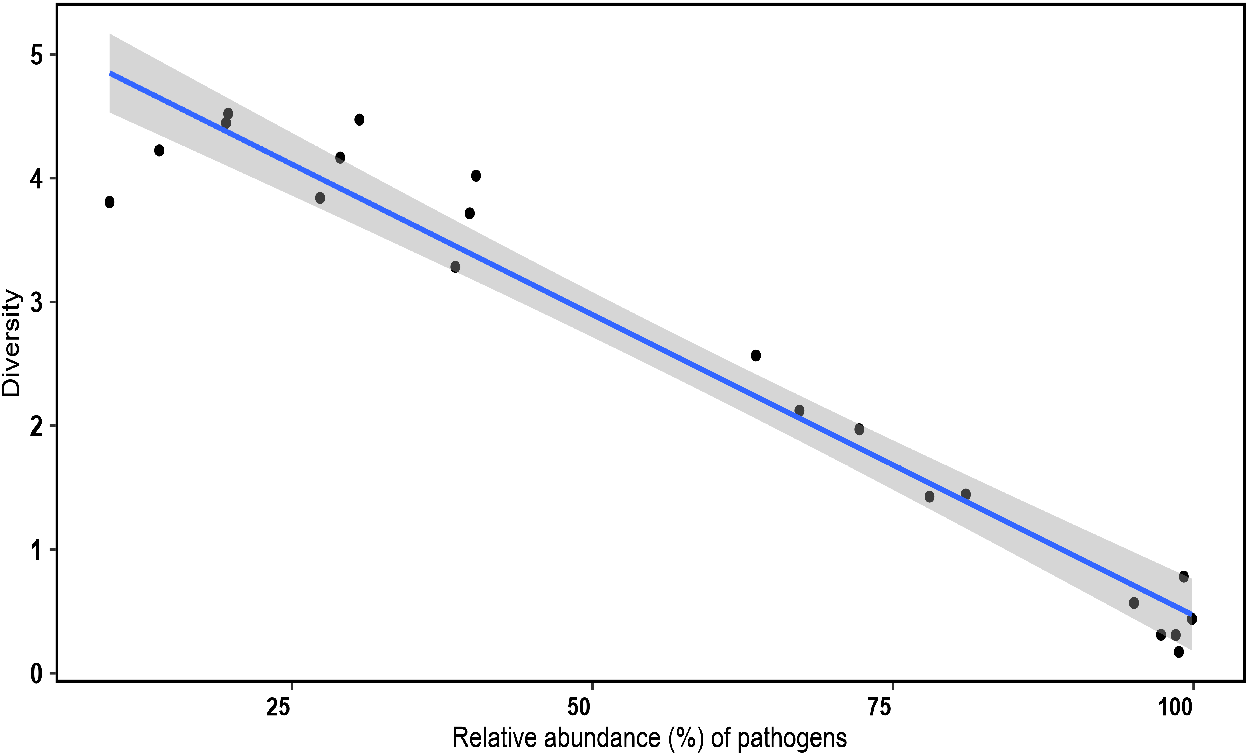
Pearson correlation between the Shannon diversity index and the relative abundance of pathogenic species identified in the 21 COVID-19 patients.

Dickson and collaborators (2014, 2015) (*6, 21*) report that, while in healthy lungs the immigration and elimination processes are more determinant for microbial composition, in advanced lung diseases, pulmonary pathogenesis is a consequence of respiratory dysbiosis. Hyperventilation accelerates the influx of air-borne microbes and microbial elimination is reduced by bronchoconstriction and impaired mucociliary clearance. This condition provides a nutrient-rich growth medium and decreased oxygen concentration, influencing bacterial reproduction. In addition, these authors propose that the response to inflammation and endothelial and epithelial injury increases vascular permeability, promoting the escape of nutrient-rich fluid into the alveolar compartment, restoring the nutrient supply and selectively favoring the growth of specific lung pathogens. Our results suggest that COVID-19 reduces microbial diversity and increases the proportion of pathogens, similar to other acute and chronic lung diseases (*22*).

Almost 75% of the microbiota present in patients was composed of bacteria whose diversity was inferred from the fifty most prevalent species (Figure 2 and Table 2). Most respiratory pathogens were composed of *Acinetobacter baumannii* (corresponding to 16.5% of all bacterial species), *Streptococcus pneumoniae* (8.8%), *Pseudomonas aeruginosa* (3.6%) and *Staphylococcus aureus* (3.4%). Nosocomial pathogens mainly included fungi *Candida albicans* and *Candida tropicalis*, representing 11.2% and 8.1% of Eukarya domain, as well as the bacteria *Enterococcus faecalis* (1.5%) *and Escherichia coli* (0.5%).

**Figure 2.**
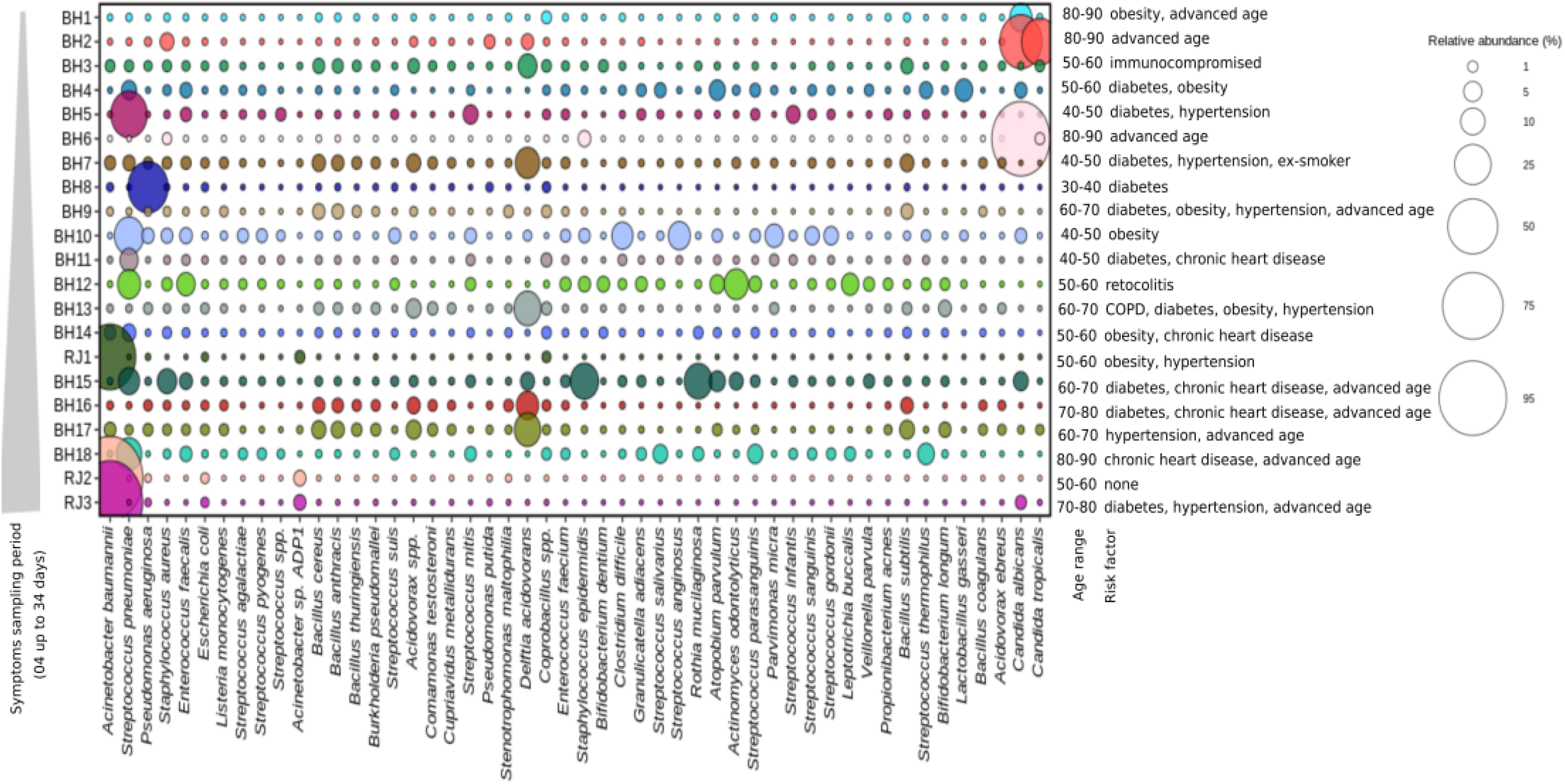
Relative abundance of the 50 most prevalent species identified in the 21 COVID-19 patients. The characteristics of the individuals analyzed, such as age range, comorbidities and time (days) from onset of symptoms until sample collection were represented.

Furthermore, the patients analyzed in our work showed a diversity of other pathogenic microorganisms, such as the uncommon *Listeria monocytogenes, Streptococcus* (*S. agalactiae* and *S. pyogenes*), sporulating *Bacillus* (*B. anthracis* and *B. thuringiensis*) and *Acidovorax spp*., although less frequently (Figure 2). Curiously, the opportunistic microorganisms *Delftia acidovorans* and *Coprobacillus spp*. were found with an abundance of almost 6% and 2%, respectively. *Delftia acidovorans* is usually not pathogenic, but catheter-related bacteremia and cases of pneumonia with lung cavity formation has been observed (*23, 24*).

As expected, an elevated occurrence of the bacterial respiratory pathogen *Streptococcus pneumoniae* was found independent of the comorbidity, while in patients without comorbidity the occurrence of respiratory and nosocomial pathogens was drastically reduced, with exception of *Candida albicans* and *Candida tropicalis* (Figure 2 and Table 2). In contrast, *Acinetobacter baumannii* was significantly increased in patients with higher SARS-CoV2 infection period (RJ2 and RJ3 samples). No significant difference or direct association was observed between pathogen abundance and comorbidity type. However, the microbiota here identified reinforce the severity of infection under weakened health conditions.

Additionally, a large diversity of the oropharynx commensal species was identified, such as *Rothia mucilaginosa, Parvimonas micra, Actinomyces odontolyticus*, and *Streptococcus anginosus* (Table 2). *R. mucilaginosa* has been associated to pneumonia in patients with chronic obstructive pulmonary disease (COPD) (*25*). Furthermore, an association of co-infection by *A. odontolyticus and P. micra* or *Streptococcus spp*. to lung abscess and acute respiratory failure was reported (*26, 27*). Although respiratory disease caused by these commensals is a rare event, their presence in individuals with compromised immune responses should not be neglected.

The bacterial pathogens identified in this study, particularly *Acinetobacter baumannii, Streptococcus pneumoniae, Pseudomonas aeruginosa* and *Staphylococcus aureus*, may promote virulence and evade the host immune response by biofilm production, induction of hemolysins, pneumolysin, phospholipases, iron acquisition factors, cytokines, adhesins, and complement system resistance, besides other virulence factors (*28 - 30*).

In pulmonary and catheter-associated infections, biofilm formation can be favored by microbial interaction in endotracheal tubes and mechanical-ventilation apparatus, resulting in a diverse microbial complex and enhanced antimicrobial resistance (*31,32*). Studies demonstrated that *A. baumannii, Klebsiella pneumoniae*, and *Enterococcus faecium* are also frequently recovered from endotracheal tubes (*33-35*). In addition, in clinical isolates with persistent infections, *P. aeruginosa, S. aureus* and *C. albicans* are the predominant biofilm-producers and species with more biofilm formation capacity are frequently observed to be multidrug-resistant (*36-38*). For some of these microorganisms, the increase of biofilm, production of bacterial virulence factors and antibiotic resistance are related to enrichment of macrophage secretory products in culture and oxygen-limiting conditions (*39, 40*). In *S. aureus*, for example, biofilm is related to accumulation of activated macrophages exhibiting anti-inflammatory and pro-fibrotic properties (*41*). Moreover, it was observed that, when in biofilm, bacterial species are more resistant to the immune response, evading neutrophil mediated phagocytosis (*42*).

Hemolysin, an important virulence factor of *S. aureus, E. coli*, and *Enterococcus faecalis* (*43, 44*), is a protein that causes erythrocyte lyse and disrupts the cell membrane, contributing to lung injury and pneumonia. In *Streptococcus pneumoniae*, when pulmonary surfactant is deficient, there is an expression of hemolysin, which is correlated with lung epithelial cell injury and induction of interleukin (IL)–8 (*45*).

Similar to hemolysin, pneumolysin is a pore-forming toxin, and most of its isoforms exhibit hemolytic activity. This protein is crucial for virulence and chronic bacteraemia of *Streptococcus pneumoniae*, and together with polysaccharide capsule proteins and adhesins reduce the *S. pneumoniae* entrapment in the pulmonary mucus, permitting adherence and inflammatory responses and activating complement and apoptosis (*46*).

For *Candida* species identified in this work, phospholipase and biofilm are described as the major virulence factors (*47, 48*). Phospholipases possess an important role in lung diseases, promoting change in membrane composition, stimulating chemokines and cytokines, and altering gas exchange and as a lung surfactant (*49*).

Hemolysin, phospholipase and biofilm are some of the many pathogenicity factors in *Acinetobacter baumannii*. Successful fitness of this bacteria, however, is a consequence of a large virulence repertoire, also composed by outer membrane proteins, secretion systems, surface adhesins, glycoconjugates and iron-chelating activities, which can explain its multi-resistance to antibiotics in nosocomial infections (*28*).

Biofilm-based infections are significantly less susceptible to antimicrobial agents and their treatment is extremely difficult. According to Sanchez et al., 2013 (*37*), the investigation of biofilm forming capacity in patients with diverse infective sources revealed that *Staphylococcus aureus, Acinetobacter baumannii, Pseudomonas aeruginosa, Klebsiella pneumoniae*, and *Escherichia coli* are strong biofilm producers, at levels greater than or equal to *Staphylococcus epidermidis*, a positive biofilm producing strain. The greatest number of biofilm producing strains corresponded to *P. aeruginosa* and *S. aureus* species.

Interestingly, the metabolic profile obtained in our study correlated to virulence factors of the species described, particularly biofilm (Tables 3-6). Within Central Carbohydrate Metabolism (Table 3), the TCA cycle was the predominant pathway (18%), with abundance of dihydrolipoamide dehydrogenase enzyme (Table 4). According to Lu and authors (2019) (*50*), alteration in the metabolites from the TCA cycle, amino acid metabolism, and glycerolipid pathways were observed during biofilm production. Furthermore, characterization of dihydrolipoamide dehydrogenase from *Streptococcus pneumoniae* showed that this enzyme plays an important role in pneumococcal infection, being necessary for the survival and capsular polysaccharide production of pneumococci within the host (*51*).

**Table 3.** Functional pathway of Central Carbohydrate Metabolism, related to virulence factors reported for the species identified in this work.

| Level 1 | Level 2 | Level 3 | Frequency |
| --- | --- | --- | --- |
| Carbohydrates | Central carbohydrate metabolism | TCA_Cycle | 18.0406 |
| Carbohydrates | Central carbohydrate metabolism | Pentose_phosphate_pathway | 13.0674 |
| Carbohydrates | Central carbohydrate metabolism | Pyruvate_metabolism_I:_anaplerotic_reactions,_PEP | 12.7037 |
| Carbohydrates | Central carbohydrate metabolism | Pyruvate_metabolism_II:_acetyl-CoA,_acetogenesis_from_pyruvate | 10.5659 |
| Carbohydrates | Central carbohydrate metabolism | Dehydrogenase_complexes | 8.8438 |
| Carbohydrates | Central carbohydrate metabolism | Glycolysis_and_Gluconeogenesis_including_Archaeal_enzymes | 7.6641 |
| Carbohydrates | Central carbohydrate metabolism | Glycolysis_and_Gluconeogenesis | 7.2847 |
| Carbohydrates | Central carbohydrate metabolism | Pyruvate_Alanine_Serine_Interconversions | 6.9584 |
| Carbohydrates | Central carbohydrate metabolism | Entner-Doudoroff_Pathway | 4.7727 |
| Carbohydrates | Central carbohydrate metabolism | HPr_kinase_and_hprK_operon_in_Gram-positive_organisms | 2.7760 |
| Carbohydrates | Central carbohydrate metabolism | Glycolate,_glyoxylate_interconversions | 1.8322 |
| Carbohydrates | Central carbohydrate metabolism | Pyruvate:ferredoxin_oxidoreductase | 1.6865 |
| Carbohydrates | Central carbohydrate metabolism | Glyoxylate_bypass | 1.3451 |
| Carbohydrates | Central carbohydrate metabolism | Peripheral_Glucose_Catabolism_Pathways | 1.2182 |
| Carbohydrates | Central carbohydrate metabolism | Dihydroxyacetone_kinases | 0.8161 |
| Carbohydrates | Central carbohydrate metabolism | Methylglyoxal_Metabolism | 0.4227 |
| Carbohydrates | Central carbohydrate metabolism | Ethylmalonyl-CoA_pathway_of_C2_assimilation | 0.0018 |

**Table 4.** Functional profile of TCA_Cycle pathway, related to virulence factors reported for the species identified in this work.

| Level 1 | Level 2 | Level 3 | Function | Total abundance | BH1 | BH2 | BH3 | BH4 | BH5 | BH6 | BH7 | BH8 | BH9 | BH10 | BH11 | BH12 | BH13 | BH14 | BH15 | BH16 | BH17 | BH18 | RJ1 | RJ2 | RJ3 |
| --- | --- | --- | --- | --- | --- | --- | --- | --- | --- | --- | --- | --- | --- | --- | --- | --- | --- | --- | --- | --- | --- | --- | --- | --- | --- |
| Carbohydrates | Central carbohydrate metabolism | TCA_Cycle | Dihydrolipoamide dehydrogenase (EC 1.8.1.4) | 34640 | 5 | 820 | 250 | 485 | 275 | 305 | 14155 | 175 | 870 | 0 | 35 | 3390 | 50 | 30 | 2320 | 80 | 280 | 200 | 65 | 6120 | 4730 |
| Carbohydrates | Central carbohydrate metabolism | TCA_Cycle | Succinate dehydrogenase flavoprotein subunit (EC 1.3.99.1) | 19476 | 18 | 582 | 492 | 144 | 60 | 153 | 1863 | 231 | 552 | 36 | 6 | 1560 | 102 | 30 | 1554 | 177 | 480 | 12 | 81 | 6591 | 4752 |
| Carbohydrates | Central carbohydrate metabolism | TCA_Cycle | Isocitrate dehydrogenase [NADP] (EC 1.1.1.42) | 18878 | 8 | 654 | 306 | 62 | 16 | 242 | 2112 | 156 | 228 | 44 | 10 | 184 | 108 | 22 | 338 | 196 | 360 | 28 | 88 | 7842 | 5874 |
| Carbohydrates | Central carbohydrate metabolism | TCA_Cycle | 2-oxoglutarate dehydrogenase E1 component (EC 1.2.4.2) | 15954 | 12 | 586 | 332 | 40 | 22 | 192 | 1342 | 220 | 198 | 26 | 16 | 1102 | 86 | 24 | 1246 | 172 | 366 | 28 | 82 | 5582 | 4280 |
| Carbohydrates | Central carbohydrate metabolism | TCA_Cycle | Citrate synthase (si) (EC 2.3.3.1) | 13758 | 6 | 516 | 309 | 108 | 18 | 210 | 3129 | 177 | 225 | 36 | 12 | 933 | 129 | 12 | 405 | 183 | 366 | 63 | 51 | 3972 | 2898 |
| Carbohydrates | Central carbohydrate metabolism | TCA_Cycle | Succinate dehydrogenase iron-sulfur protein (EC 1.3.99.1) | 13410 | 5 | 285 | 305 | 110 | 25 | 165 | 1360 | 110 | 370 | 40 | 25 | 1145 | 50 | 15 | 1200 | 125 | 370 | 5 | 60 | 4405 | 3235 |
| Carbohydrates | Central carbohydrate metabolism | TCA_Cycle | Fumarate hydratase class II (EC 4.2.1.2) | 6109 | 13 | 204 | 122 | 49 | 5 | 103 | 1920 | 66 | 162 | 10 | 9 | 194 | 47 | 8 | 434 | 32 | 159 | 5 | 10 | 1401 | 1156 |
| Carbohydrates | Central carbohydrate metabolism | TCA_Cycle | Malate:quinone oxidoreductase (EC 1.1.99.16) | 4162 | 0 | 74 | 26 | 5 | 0 | 36 | 676 | 10 | 109 | 0 | 0 | 66 | 5 | 0 | 473 | 16 | 23 | 0 | 21 | 1509 | 1113 |
| Carbohydrates | Central carbohydrate metabolism | TCA_Cycle | Dihydrolipoamide dehydrogenase of pyruvate dehydrogenase complex (EC 1.8.1.4) | 2532 | 0 | 98 | 218 | 28 | 18 | 40 | 770 | 132 | 18 | 40 | 10 | 264 | 60 | 6 | 430 | 138 | 244 | 8 | 0 | 10 | 0 |
| Carbohydrates | Central carbohydrate metabolism | TCA_Cycle | Isocitrate dehydrogenase [NAD] (EC 1.1.1.41) | 686 | 5 | 24 | 0 | 0 | 0 | 12 | 418 | 6 | 0 | 0 | 0 | 182 | 0 | 0 | 20 | 5 | 0 | 6 | 0 | 8 | 0 |
| Carbohydrates | Central carbohydrate metabolism | TCA_Cycle | Isocitrate dehydrogenase [NAD] subunit I, mitochondrial precursor (EC | 227 | 0 | 112 | 0 | 0 | 0 | 90 | 12 | 0 | 0 | 0 | 0 | 0 | 0 | 0 | 0 | 0 | 0 | 0 | 0 | 0 | 13 |
|  |  |  | 1.1.1.41) |  |  |  |  |  |  |  |  |  |  |  |  |  |  |  |  |  |  |  |  |  |  |
| Carbohydrates | Central carbohydrate metabolism | TCA_Cycle | Isocitrate dehydrogenase [NAD] subunit II, mitochondrial precursor (EC 1.1.1.41) | 61 | 0 | 26 | 0 | 0 | 0 | 22 | 8 | 0 | 0 | 0 | 0 | 0 | 0 | 0 | 0 | 0 | 0 | 0 | 0 | 0 | 5 |
| Carbohydrates | Central carbohydrate metabolism | TCA_Cycle | Succinyl-CoA ligase [GDP-forming] alpha chain (EC 6.2.1.4) | 42 | 0 | 24 | 0 | 0 | 0 | 18 | 0 | 0 | 0 | 0 | 0 | 0 | 0 | 0 | 0 | 0 | 0 | 0 | 0 | 0 | 0 |
| Carbohydrates | Central carbohydrate metabolism | TCA_Cycle | Succinyl-CoA ligase [GDP-forming] beta chain (EC 6.2.1.4) | 37 | 0 | 21 | 0 | 0 | 0 | 16 | 0 | 0 | 0 | 0 | 0 | 0 | 0 | 0 | 0 | 0 | 0 | 0 | 0 | 0 | 0 |
| Carbohydrates | Central carbohydrate metabolism | TCA_Cycle | Citrate lyase gamma chain, acyl carrier protein (EC 4.1.3.6) | 28 | 0 | 0 | 0 | 0 | 0 | 0 | 12 | 0 | 0 | 0 | 0 | 11 | 0 | 0 | 5 | 0 | 0 | 0 | 0 | 0 | 0 |

Besides enzymes associated with biofilm formation, we identified adhesins, permeases for antibiotic export, resistance and toxins, and superantigens belong to the Virulence, Disease and Defense functional category (Table 5). The most abundant adhesins are fibronectin/fibrinogen-binding proteins, although high numbers of sortases, related to attachment of specific proteins to the cell wall, were also found. Toxins and superantigens are composed of the streptolysin S locus, which produces the beta-haemolytic phenotype. Streptolysin S causes impaired phagocytic clearance and promotes epithelial cell cytotoxicity, enhancing the effects of M protein and streptolysin O (*52*). In the Membrane Transport category, protein secretion system Types I, V, and VI are highlighted, with abundance of exoprotein involved in heme utilization, cytolysin activator and adhesin (Table 6).

**Table 5.**
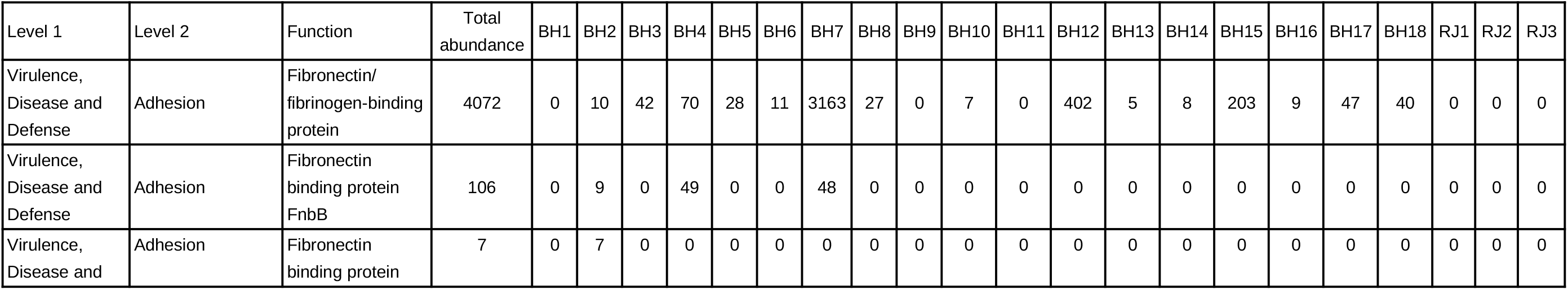
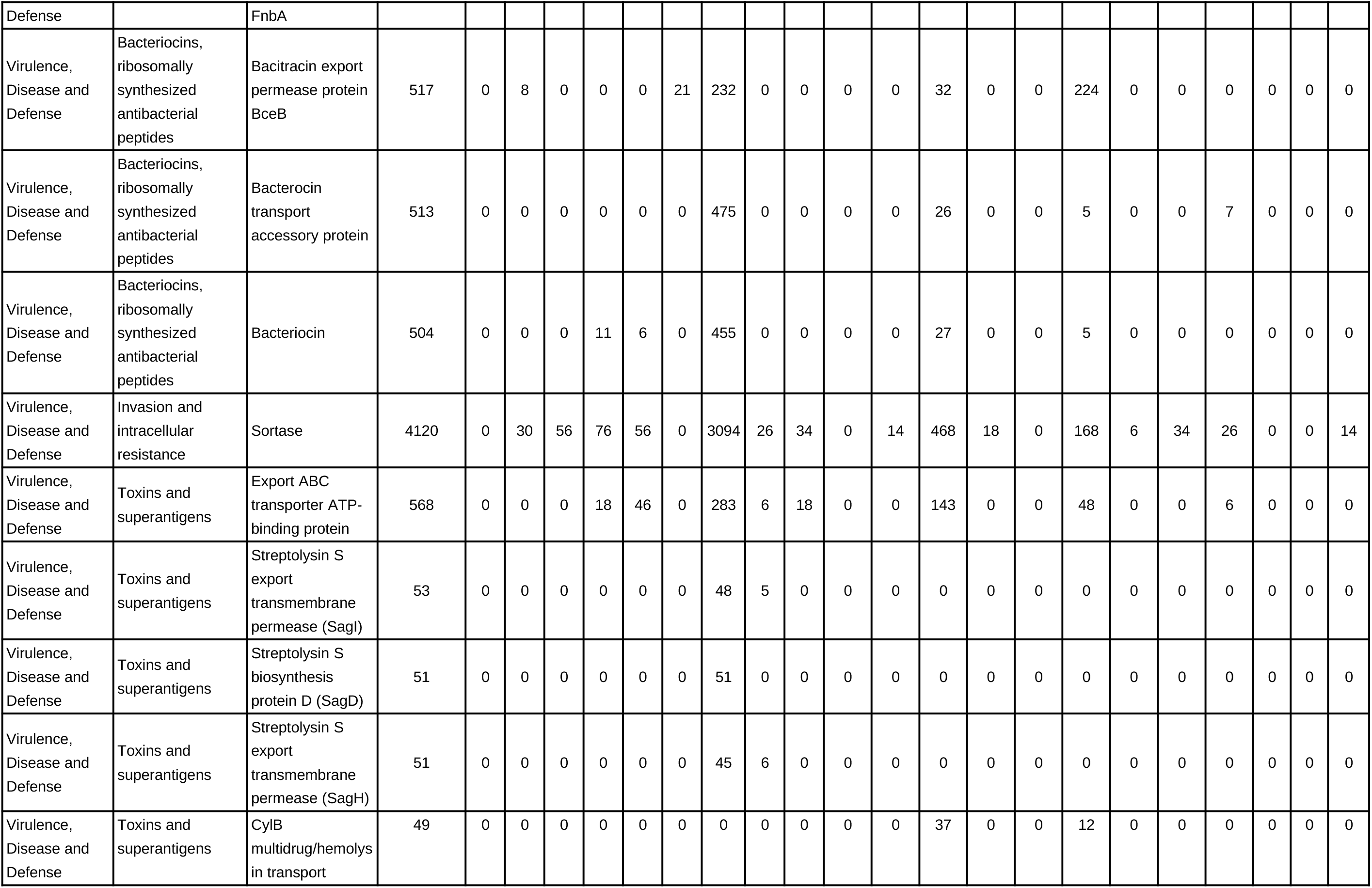
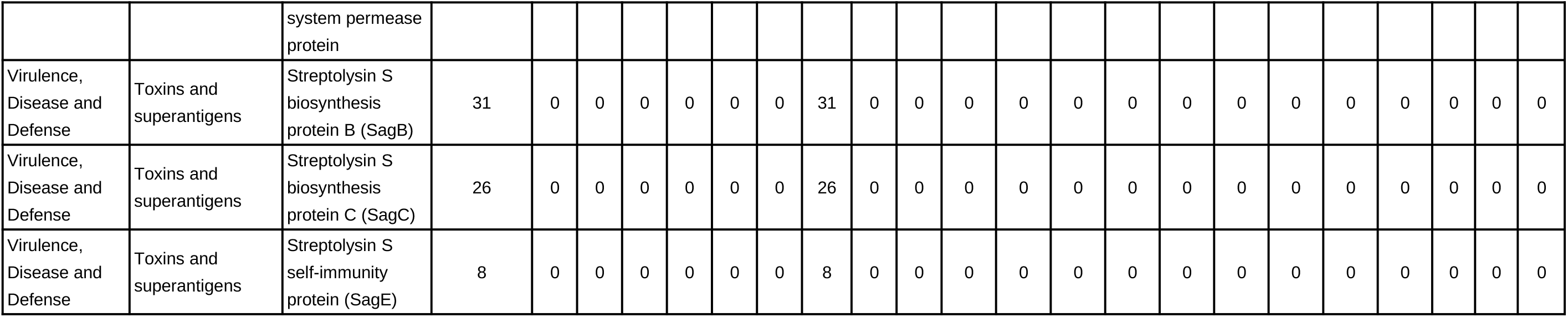
Functional profile of Virulence, Disease and Defense pathway, related to virulence factors reported for the species identified in this work.

**Table 6.**
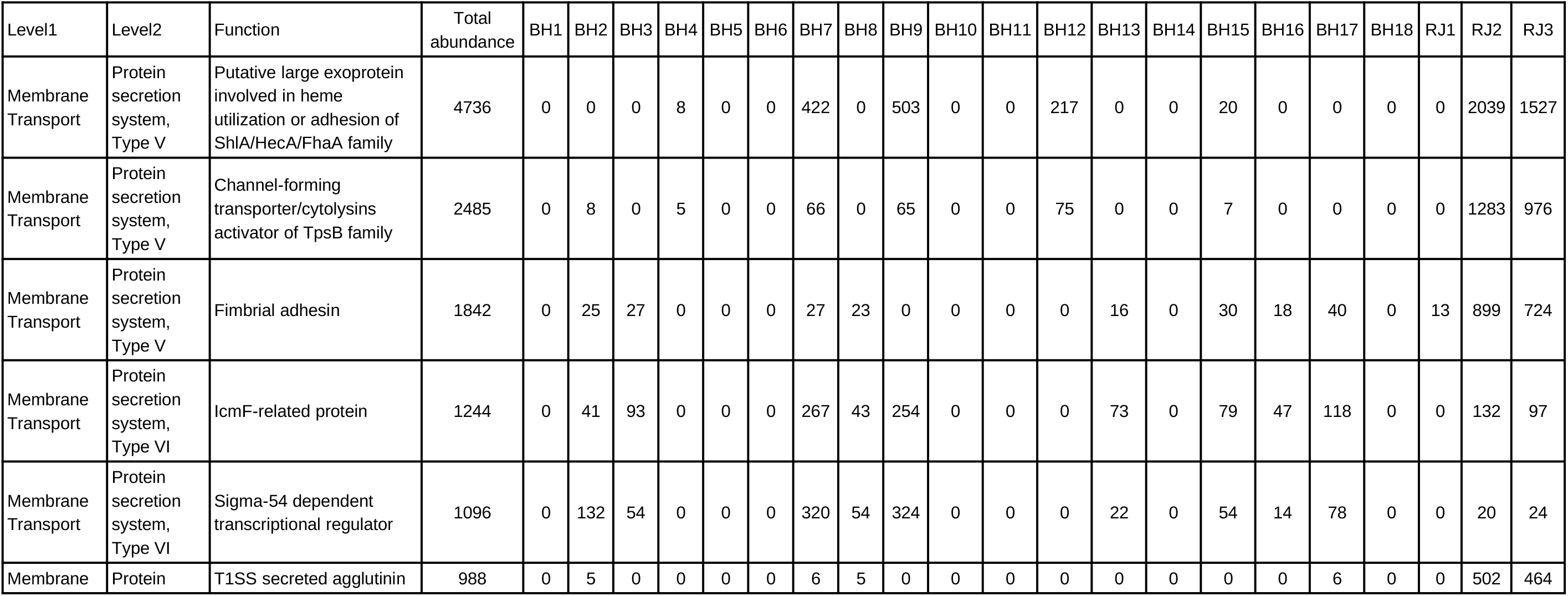
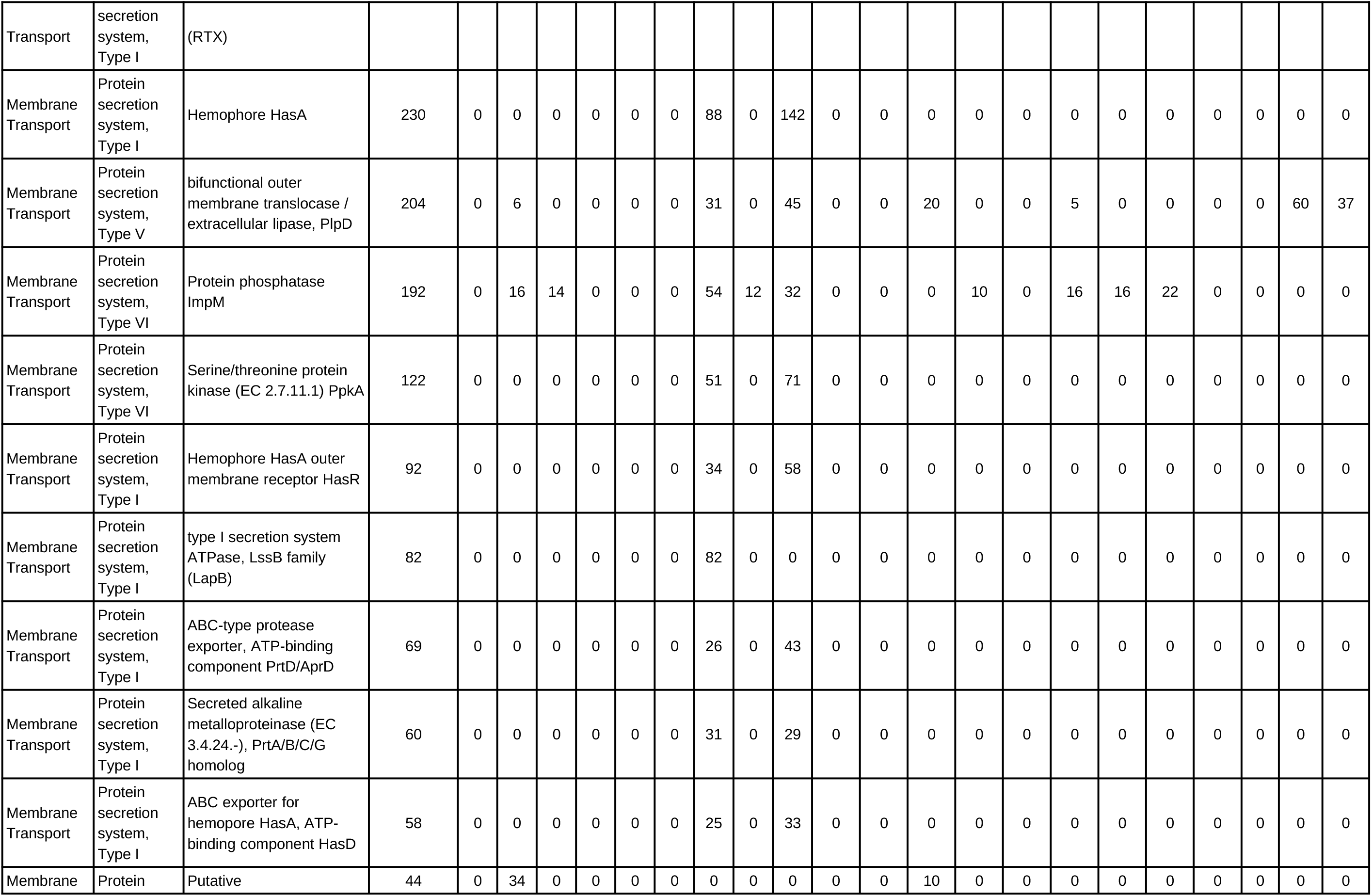
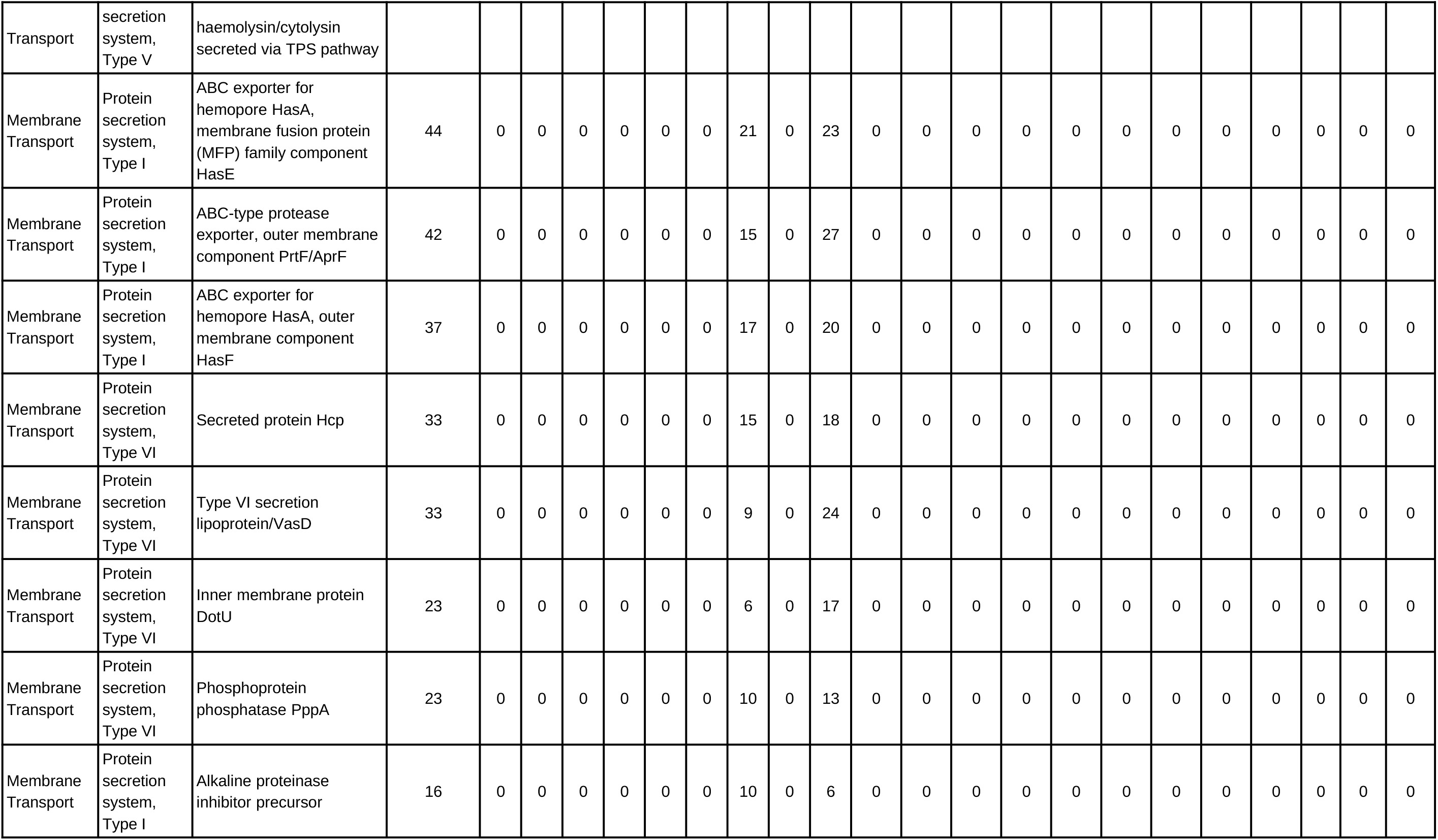
Functional profile of Membrane Transport pathway, related to virulence factors reported for the species identified in this work.

The scenario highlights the high occurrence of potentially virulent pathogens, which can contribute to severity of lung infection in Coronavirus Disease 2019 patients admitted to intensive care units. Moreover, the prevalence of respiratory pathogens observed in COVID-19 patients analyzed can be orchestrated mainly by the complex and distinct immune events in response to lung damage, as well as by the ecological model proposed by Dickson and collaborators (2014, 2015) (*26, 25*), which indicate that several lung diseases can alter the growth of local microbiota, leading to an increase in bacterial abundance. A cellular model to explain the bacterial species described in our study correlated to processes that aggravate COVID-19 is presented (Figure 3).

**Figure 3.**
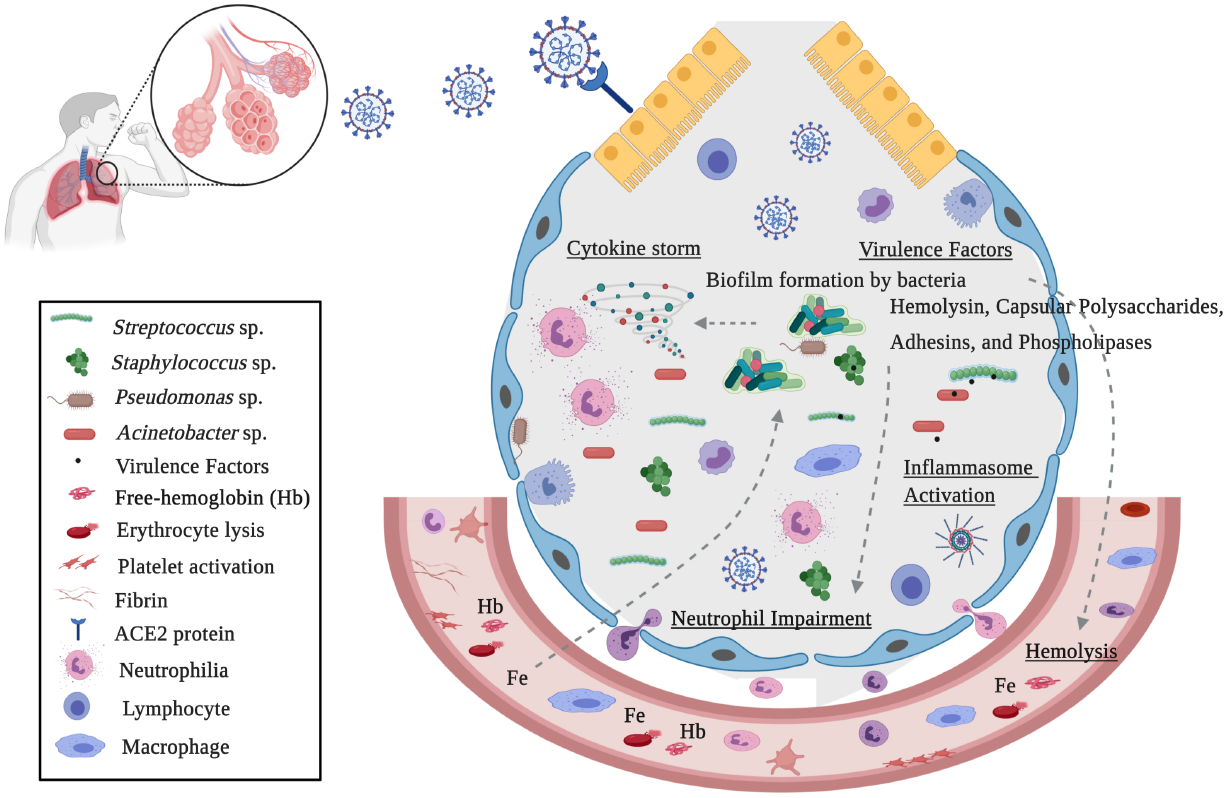
Model for an interplay of the bacteria found in COVID-19 patients, virulence factors and free-iron in immune response and disease severity. In severe lung diseases, hyperventilation contributes to microbial influx and reduces the elimination of microorganisms. Tecidual injury promotes a rich environment for growth of specific lung pathogens into the alveolar compartment. Bacterial pathogens evade the host immune response by biofilm production, induction of hemolysins, iron acquisition factors, adhesins, and other virulence factors. The SARS-CoV-2 and bacterial pathogens promote erythrocyte lysis, causing heme(Hb)/iron(Fe) liberation. Heme uptake is required for bacterial host colonization and increased production of virulence factors. Bacterial virulence factors, as well as free-heme, activates the inflammasome, contributing to neutrophil inflammation, cytokine elevation and pathogenesis processes. This figure was generated using the BioRender (available at https://biorender.com/).

Impairment in neutrophil function, macrophage depletion and excessive inflammation has been reported as an important factor in the progression of lower respiratory tract pathologies (*53*). It was demonstrated that alveolar macrophage depletion could facilitate the bacterial infection by the establishment of a niche for secondary *Streptococcus pneumoniae* infection, altering cellular innate immunity and resulting in lethal pneumonia (*54*). Additionally, neutrophilia plays an important role in bacterial pulmonary diseases, including those caused by *Streptococcus pneumoniae, Klebsiella pneumoniae, Haemophilus influenzae*, and *Staphylococcus aureus* (*55*). Interestingly, Sodhi and collaborators (2019) (*56*) demonstrated that impaired neutrophil and innate immune response might be mediated by variations in the activity of angiotensin-converting enzyme 2 (ACE2). In bacterial lung infection, when pulmonary active ACE2 levels vary dynamically, neutrophil influx increases, compromising the host-defense capability and heightened inflammatory response, with subsequent elevation of infection severity and mortality

Similar to Coronavirus Disease 2019, patients with chronic obstructive pulmonary disease (COPD) show vascular permeabilization, platelet activation, thrombosis, adhesion molecule expression, leukocyte recruitment, and complement activation, which are also observed in injury-derived free heme (*57*). Su and authors (2020)(*58*) observed increased heme and heme oxygenase (HO)-1 in the blood system of severe COVID-19 patients. Moreover, an additional role of hemoglobinopathy, hypoxia and hyperferritinemia has been proposed in worsening COVID-19 infection (*59*).

Increase of cell-free hemoglobin (Hb) and HO-1 expression in lung tissue and bronchoalveolar lavage fluid has been correlated to COPD and acute respiratory distress syndrome (ARDS) severity (*60*). Simon and authors (2009)(*61*) demonstrated an ACE-like activity of Hb in the conversion of angiotensin I to its active metabolites induced by ferrous- and ferryl-Hb, with a possible effect on vasoconstriction. Moreover, intratracheal administration of heme in mice led to alveolar-capillary barrier dysfunction and increased alveolar permeability, contributing to acute lung and ARDS (*62*). Curiously, heme uptake by pathogens plays an important role during bacterial infection. These microorganisms induce erythrocyte hemolysis caused by pore-forming toxins, such as hemolysin and phospholipases. Free-heme and iron acquired from erythrocyte disruption is required for invasion, growth and successful pathogen colonization in the host (*63*). In addition, Dutra and collaborators (2014)(*64*) showed that heme promotes the processing of caspase-1, inflammasome activation and IL-1β secretion by macrophages that participate in the immune response induced by hemolysis, contributing to inflammation and pathogenesis. Strategies for interfering in inflammasome activation by bacterial pathogens have evolved (*65*), and together with SARS-Cov2, may contribute to a hyperactivated inflammatory response (*66*).

Interdependence between iron acquisition and virulence factors for bacterial pathogenesis have been demonstrated. In *Streptococcus pneumoniae*, iron is a critical factor for regulation of several enzymes. During pneumonia and bacteremia, the transcription of hemolysin, toxins, cell wall hydrolases and biofilm formation are increased in a positive Fe-dependent regulation (*67*). For *Pseudomonas aeruginosa*, high iron concentrations are correlated to stimulation of aggregation, adhesion and biofilm formation, in a positive-modulation manner (*68*). The acquisition of heme-iron bound to erythrocyte hemoglobin is the preferred source of iron during *S. aureus* infection initiation, as well as for its proliferation. In the presence of heme, regulatory systems induced exotoxins such as cytolysins, proteases, and other virulence factors, promote Staphylococcal cellular adherence to host cells and resistance to neutrophil killing. Furthermore, iron and oxygen conditions have been shown to influence the TCA cycle activity of *Staphylococcus aureus* (69 - *71*). Free-iron uptake by ferritin and haemolytic activity is employed by *Candida albicans*. Interestingly, it has been speculated that co-habitation of *C. albicans* with both commensal and pathogenic bacteria, in a synergistic interaction, enables *C. albicans* to obtain iron more easily (*72*).

In conclusion, the presence of respiratory pathogens, nosocomial bacteria and opportunistic microorganisms during colonization of the host, secrete an extensive diversity of virulence factors. These molecules favor hemolysis, cytokine activation and inflammation. The presence of free-iron exacerbates bacterial pathogenesis in cyclic connection with the worsening of the immune response, favoring the resistance of bacterial species. These findings provide intriguing insights into SARS-CoV2-bacterial interactions contributing to COVID-19 severity.

## Data Availability

NGS data generated in our study is publicly available in SRA-NCBI (www.ncbi.nlm.nih.gov/sra), Bioproject accession PRJNA683652.

## Acknowledgments

This work was developed under the frameworks of Corona-ômica-RJ (FAPERJ = E-26/210.179/2020) and Rede Corona-ômica BR MCTI/FINEP (FINEP = 01.20.0029.000462/20, CNPq = 404096/2020-4). A.T.R.V. is supported by Conselho Nacional de Desenvolvimento Científico e Tecnológico - CNPq (303170/2017-4) and FAPERJ (E-26/202.903/20). R.S.A. is supported by CNPQ 312688/2017-2, 439119/2018-9; FAPERJ 202.922/2018 .C.C.C is supported by FAPERJ (E-26/202.791/2019) and S.P.C.B is supported by FAPERJ (E-26/010.000168/20).

## Author Bio

F.M.C is currently a postdoctoral fellow in the Laboratory of Bioinformatics (LABINFO) of the National Laboratory of Scientific Computing (LNCC). Research interests comprise metagenomic (clinical and environmental) and comparative genomics.

